# Incidence and Outcomes of Atrial Fibrillation and Systolic Dysfunction in Patients Receiving Mavacamten for Obstructive Hypertrophic Cardiomyopathy: A Multicenter Study

**DOI:** 10.1101/2025.09.15.25335783

**Authors:** Olives Nguyen, Jack Wiedrick, Daniele Massera, Elizabeth Adlestein, Sumar Frejat, Matteo Castrichini, Said Alsidawi, John R. Giudicessi, Jeffrey B. Geske, Richard T. Carrick, Jose Madrazo, Nicole Dellise, Mark A. Zenker, Thomas A. Boyle, Nosheen Reza, Anjali Tiku Owens, David S. Frankel, Prabhjot Hundal, Jamil Tajik, Patrycja Galazka, Myra Lewontin, Michael Ayers, Timothy Wong, Michael Flanagan, Sumeet Singh Mitter, Arjun Kanwal, Ozlem Bilen, Sarah Baghdadi, Hirak Shah, Jared Kvapil, Paola Roldan, Loren Berenbom, Jill Jesurum, Denise Tootill, Alice Siqueira-Benzow, Mariko Harper, Danish Saleh, Lubna Choudhury, Isabela Valenta, Melissa Lang, Dermot M. Phelan, Dmitry Prizand, Neal Lakdawala, Carolyn Y. Ho, Lusha W. Liang, Shepard D. Weiner, Sririam Ravi, Ahmed Sami Abuzaid, Mohammed Makkiya, Jeremy S. Markowitz, Mark Sherrid, Ahmad Masri

**Affiliations:** Knight Cardiovascular Institute, Oregon Health & Science University (Portland, OR, USA); NYU Langone (New York, NY, USA); Mayo Clinic (Rochester, MN, and Phoenix, AZ, USA); Johns Hopkins University (Baltimore, MD, USA); Ascension Saint Thomas Heart West (Nashville, TN, USA); University of Pennsylvania (Philadelphia, PA, USA); Advocate Health - Aurora St. Luke’s Medical Center (Milwaukee, WI, USA); The University of Virginia (Charlottesville, VA, USA); University of Pittsburg Medical Center (Pittsburgh, PA, USA); INOVA (Springfield, VA, USA); Emory University Hospital (Atlanta, GA, USA); The University of Kansas Health System (Kansas City, Kansas, USA); Virginia Mason Franciscan Health HCM Center of Excellence (Seattle, WA, USA); Northwestern University (Chicago, Illinois, USA); Atrium Health - Sanger Heart & Vascular Institute (Charlotte, NC, USA); Brigham and Women’s Hospital (Boston, MA, USA); Columbia University Medical Center (New York, NY, USA); David Heller Center for Hypertrophic Cardiomyopathy, Providence Heart Institute (Portland, OR, USA); Alaska Heart and Vascular Institute (Anchorage, AK, USA); Virginia Commonwealth University (Richmond, VA, USA); University of Minnesota Heart Care: Genetics Clinic (Minneapolis, MN, USA)

**Author notes:** Corresponding author: Ahmad Masri, MD MS, Associate Professor of Medicine, Mail code: UHN-62, 3181 SW Sam Jackson Rd, Portland, OR 97239.

## Abstract

**Importance:** Mavacamten is highly effective in treating symptomatic obstructive hypertrophic cardiomyopathy (oHCM) and was approved for commercial use with a risk mitigation program (REMS) to monitor the impact on left ventricular systolic function (LVSD). The impact of mavacamten on atrial fibrillation (AF) occurrence is not well characterized.

**Objective:** Determine the real-world incidence of new-onset and recurrent AF among patients with HCM treated with commercial mavacamten. Secondary objectives included assessing the incidences of LVSD, heart failure (HF), and cardiogenic shock.

**Design:** A multicenter cohort study. Center-level data were aggregated for patients who received mavacamten from May 2022 through December 2024.

**Setting:** Twenty-one outpatient HCM centers in the United States.

**Participants:** Consecutive patients ≥18 years of age with oHCM were included; those with permanent AF were excluded.

**Exposures:** At least one dose of commercial mavacamten.

**Main Outcomes and Measures:** Incidence of new-onset and recurrent AF, LVSD, HF, and cardiogenic shock.

**Results:** Among 1,538 patients (median age, 66 years [95% CI, 65.9–66.1]; 57% female [95% CI, 54–59%]), 25% [95% CI, 22-27%] had prior AF. Median mavacamten exposure was 13.4 months [95% CI, 11.8–15.0].

Overall AF incidence was 13% [95% CI, 10–17%], including 5% [95% CI, 4–7%] new-onset and 39% [95% CI, 27–51%] recurrent AF in those with history of AF prior to mavacamten initiation. LVEF <50% occurred in 8% [95% CI, 6–9%], of whom 70% had AF. Symptomatic HF occurred in 1.5% [95% CI, 0.8–2.2%], cardiogenic shock in 0.6% [95% CI, 0.1–1.0%], and an overall permanent discontinuation of mavacamten in 7% [95% CI, 4–10%].

**Conclusions and Relevance:** In this multicenter cohort receiving commercial mavacamten, we identified an annual incidence of 5% new-onset AF and 39% recurrent AF, while 70% of patients who developed LVEF<50% had concurrent AF. Given this association and the morbidity that can be associated with AF, protocols for LVEF assessment and aggressive rhythm management following AF detection may be warranted to improve patient care. Further studies are needed to improve understanding of the impact of mavacamten on AF and patient outcomes.

**Key points:** *Question:* What are the real-world burden and consequences of atrial fibrillation (AF) in patients with hypertrophic cardiomyopathy treated with commercial mavacamten?

*Findings:* In a multicenter cohort of more than 1,500 patients, 5% experienced new-onset AF, while 39% of those with pre-existing AF experienced recurrence after mavacamten initiation. Left ventricular ejection fraction (LVEF) fell below 50% in 8% of patients, 70% of whom had concurrent AF.

*Meaning:* Real-world experience with mavacamten emphasizes that regular surveillance for AF is an important aspect of managing patients with HCM. Additionally, the identification of AF should prompt LVEF assessment and aggressive rhythm management, given the potential association between AF and decrease in LVEF. The observed incidence of AF on mavacamten highlights the need for further studies on potential mechanisms and for proactive surveillance and management strategies.

## Background

Atrial fibrillation (AF) is a common complication in hypertrophic cardiomyopathy (HCM)^1,2^. AF is attributed to atrial myopathy, chronically-increased left ventricular filling pressures, and diastolic dysfunction, which promote left atrial dilation, remodeling, and fibrosis^3^. Mavacamten improves symptoms, exercise tolerance, and quality of life in obstructive HCM (oHCM). In EXPLORER-HCM and over a 30-week treatment period, total AF events occurred in 6.5% of patients on mavacamten and 7% of patients on placebo^4^. However, longer-term data show a clinically-meaningful AF incidence on mavacamten despite robust relief of left ventricular outflow tract (LVOT) obstruction, including new-onset AF in 7.8% of patients in MAVA-LTE over a median of 3.2 years and 10.2% in VALOR-HCM over 2.5 years^5–9^. While left ventricular systolic dysfunction (LVSD) has been viewed as the primary complication of mavacamten therapy, AF in HCM is a morbid condition that is associated with increased thromboembolic risk, need for invasive procedures, and lifelong anticoagulation. As such, there is a need to evaluate the incidence of new and recurrent AF on mavacamten outside of the strict environment of clinical trials. In addition, the incidence of LVSD, heart failure (HF), and cardiogenic shock have not been studied in a large real-world multicenter study.

## Methods

We conducted an observational analysis of center-level data across 21 HCM centers in the United States. Patients with oHCM who were prescribed commercial mavacamten were included. Each center reported their aggregated data. Patients with permanent AF were excluded. Records were reviewed and verified by site investigators and ethics approval was obtained at each individual site in accordance with local practice. To account for site-level data and heterogeneity, we used: (1) robust per-site averages with jackknife-derived 95% confidence intervals and (2) random-effects meta-analysis, using Freeman–Tukey double arcsine transformation with Paule-Mandel estimation for between-site variance. Means and medians are presented with 95% confidence intervals.

## Results

Data from 1,538 patients were analyzed, with a mean of 73 [95% CI, 56-90] patients per site. Median age was 66 [95% CI, 65.9, 66.1] years, 1188 (83% [95% CI, 77-90%]) were White, 875 (57% [95% CI, 54-59%]) female, and 802 (51% [95% CI, 43-58%]) had a BMI ≥30. A total of 372 patients (25% [95% CI, 22-27%]) had a history of AF prior to mavacamten initiation. Patients received a median mavacamten dose of 5 mg [95% CI, 5-5] over a median of 13.4 [95% CI, 11.8, 15.0] months.

### Atrial Fibrillation

New-onset AF developed in 60 patients with an incidence of 5% [95% CI, 4-7%] following mavacamten initiation (*Figure 1A*) and a median exposure of 9.7 [95% CI, 6.9-12.4] months. AF recurred in 146 patients with prior history of AF (39% [95% CI, 27-51%]) over 7.0 [95% CI, 3.5-10.6] months of exposure (*Figure 1B*). The overall rate of AF, new and recurrent, was 13% [95% CI, 10-17%] over the study duration (*Figure 2A*).

**Figure 1.**
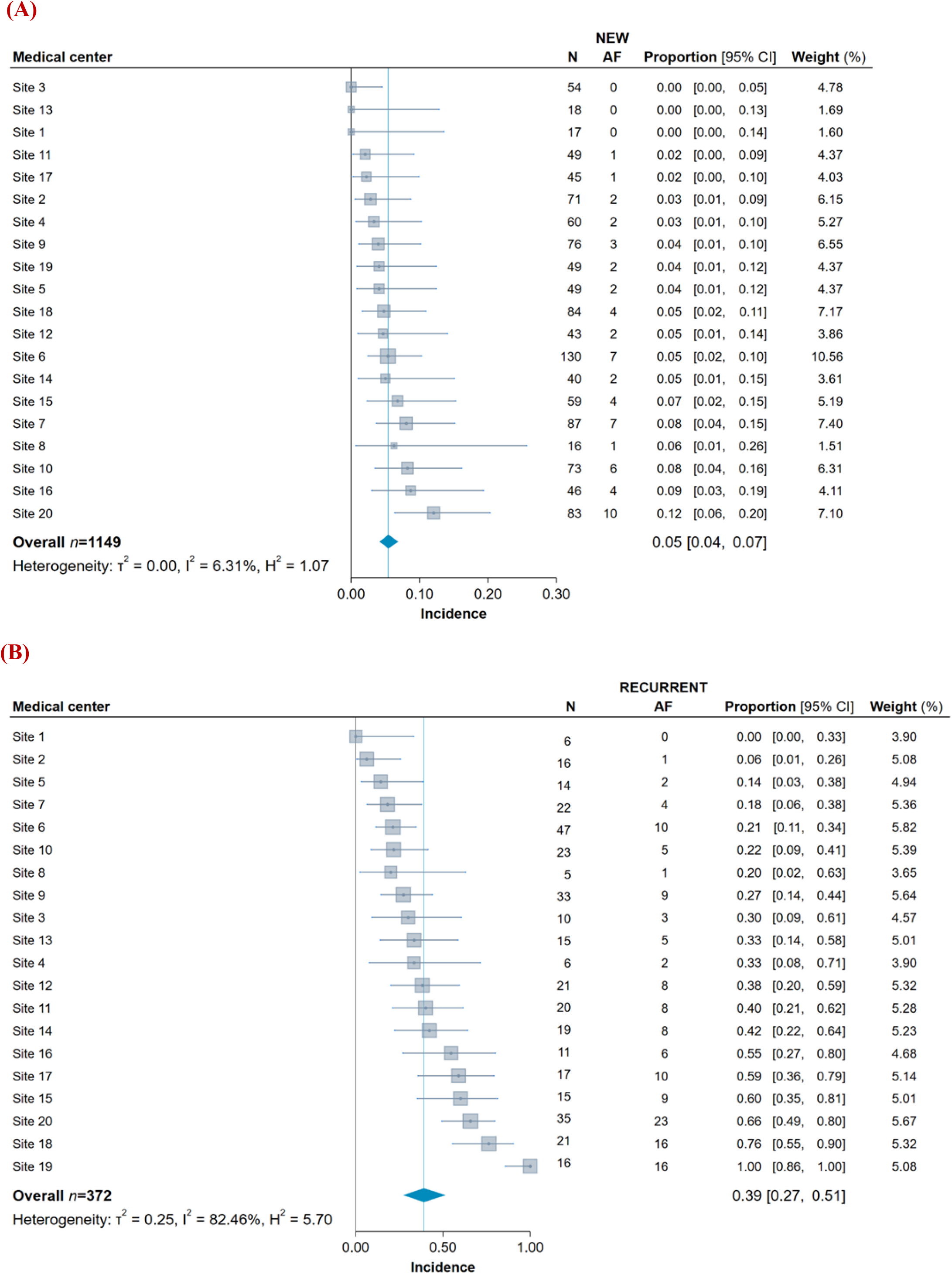
New-onset (A) and Recurrent (B) Atrial Fibrillation on Commercial Mavacamten. AF - atrial fibrillation

**Figure 2.**
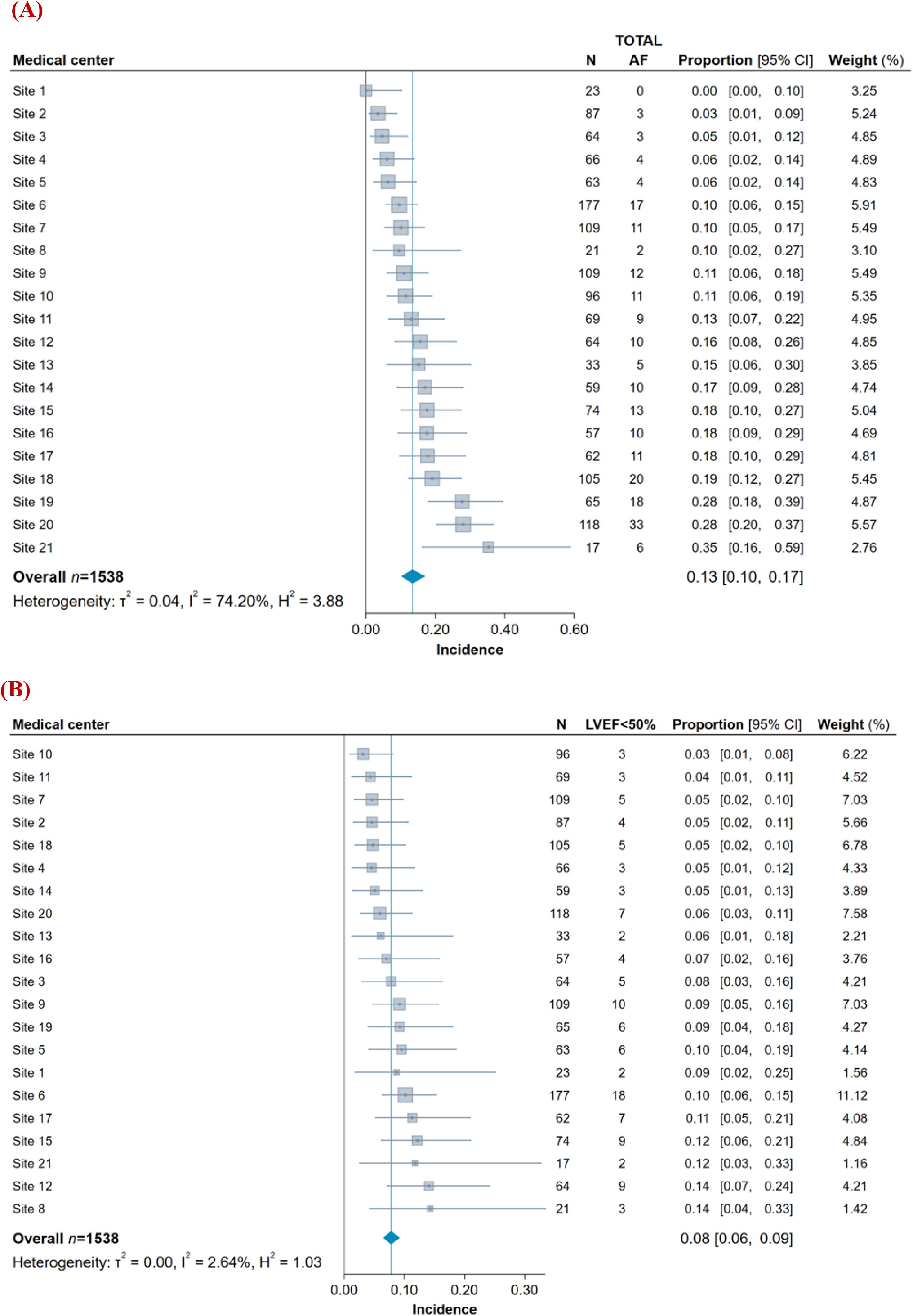
Total Atrial Fibrillation (A) and Left Ventricular Systolic Function (B) on Mavacamten. AF - atrial fibrillation, LVEF<50% - left ventricular ejection fraction less than 50%.

**Figure 3.**
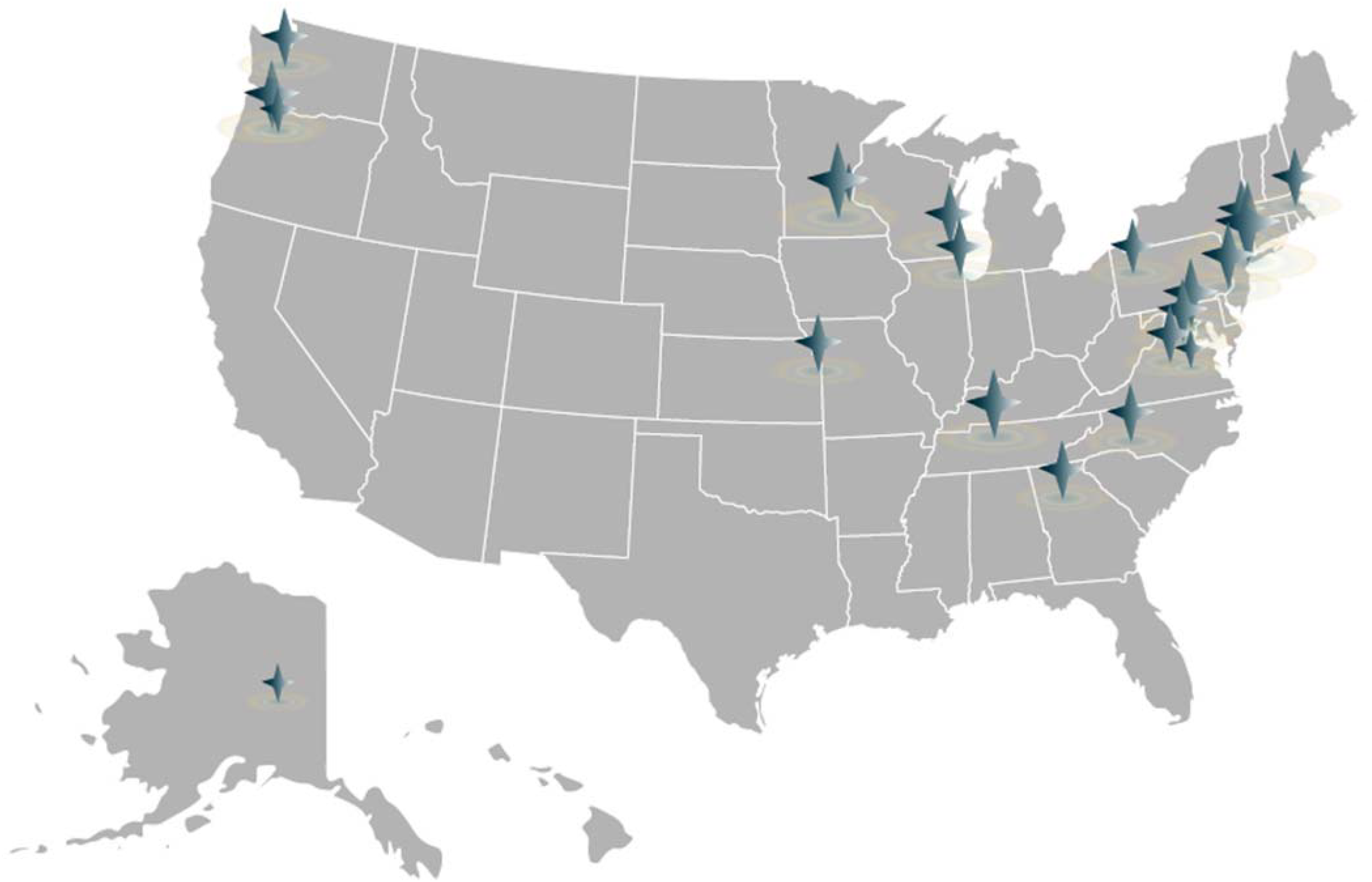
Geographical Representation of Participating Hypertrophic Cardiomyopathy Sites. Each star represents a participating site.

Among 212 patients with AF, 76 (40% [95% CI, 28–51%]) underwent direct current cardioversion (DCCV), with 30 patients (incidence: 16% [95% CI, 7–26%]) experiencing failed conversion to sinus rhythm or AF recurrence despite initial success. In contrast, 76 patients (39% [95% CI, 24–54%]) achieved sinus rhythm with medical anti-arrhythmic therapy, while 47 (22% [95% CI, 14–30%]) achieved sinus rhythm with catheter ablation.

### Left Ventricular Systolic Dysfunction

LVEF<50% occurred in 116 patients (8% [95% CI, 6-9%]) over a median of 7.8 [95% CI, 5.8-9.9] months of mavacamten therapy (*Figure 2B*). Of these patients, 79 (70% [95% CI, 45-96%]) had AF at the time of systolic dysfunction (incidence: 6% [95% CI, 3–8%]). Of those with LVEF<50%, 24 (21% [95% CI, 11–31%]) developed symptomatic HF (incidence: 1.5% [95% CI, 0.8–2.2%]). In addition, seven patients (0.6% [95% CI, 0.1–1%]) developed cardiogenic shock and five patients (0.32% [95% CI, 0.02-0.6%]) died while receiving mavacamten or shortly after discontinuing mavacamten (1 cardiogenic shock, 3 sudden cardiac death, and 1 non-cardiac death). A total of 110 patients (7% [95% CI, 3–10%]) were hospitalized due to AF, HF, or mavacamten-related complications (in the opinion of the treating physician, such as chest pain or dizziness).

Mavacamten temporary discontinuation occurred in 151 patients (12% [95% CI, 1-23%]) and permanent discontinuation in 113 patients (7% [95% CI, 4–10%]).

## Discussion

This multicenter study reports real-world experience regarding the safety of commercial mavacamten in patients with oHCM, regarding AF and HF. In more than 1,500 patients, there was a 5% incidence of new-onset AF over a median of 10 months on therapy and a 39% incidence of recurrent AF over a median of 7 months, leading to a collective incidence of AF of 13% over a median of 13 months. In addition, two-thirds of LVSD cases occurred with concomitant AF.

AF is common in HCM, with clinical trials reporting a 10–20% prevalence at enrollment despite specific eligibility criteria^5^. In EXPLORER-HCM, there was no difference in total AF events between mavacamten and placebo over 30 weeks of treatment — however, this trial’s short duration and intensive, drug-level–guided monitoring differs from real-world practice^4^. By contrast, long-term extension studies reported new-onset AF rates of 8–10%, suggesting that longer term follow-up is needed to evaluate the incidence of AF on mavacamten^8,9^. While the non-obstructive HCM population is distinct from oHCM, the ODYSSEY-HCM trial reported 14.3% of patients on mavacamten having AF as a treatment-emergent adverse event, as compared to 10% of patients on placebo over a 48-week period^10^. Mechanistically, mavacamten inhibits atrial myosin in a manner similar to ventricular myosin and mavacamten levels may vary from patient to patient based on CYP2C19 metabolizer status^11,12^. While these factors raise biologic plausibility, none establish a causal relationship between mavacamten and AF.

Mavacamten’s approval presented a pivotal moment in the treatment of patients with oHCM. Due to the need to monitor its impact on LVEF, a risk mitigation program (REMS) was imposed for commercial use^4,5,8,9^. REMS data have reported a 4.6% incidence of LVEF<50% and may represent best-case scenario reporting due to survival bias^14,15^. The current study shows double the incidence of LVSD reported in the REMS data. In addition, AF is not captured in the REMS program and is not mentioned in the FDA label since the placebo-controlled segments of trials did not show high incidence of AF with mavacamten.

Finally, we also provide new data on the incidence of cardiogenic shock occurring in 0.6% of patients prescribed mavacamten commercially.

### Limitations

The primary unavoidable limitation is the lack of a control group of comparable oHCM patients not receiving mavacamten, which precludes direct comparison with the natural history of the disease without its use. While the study included patients receiving mavacamten since its approval in 2022, the median follow up is relatively short. Further studies are needed to fully assess the impact of mavacamten on AF, LVEF, and HF. An alternative approach may be longitudinal monitoring with cardiac rhythm devices to compare atrial arrhythmia burden before and after mavacamten initiation. Additionally, we focused on safety, and as such, we did not report on the efficacy of mavacamten in this real-world cohort.

## Conclusions

In this real-world cohort of patients with oHCM treated with mavacamten, new-onset AF occurred with an incidence of 5% over a median of 13 months of follow up. Similar to clinical trials, 8% of patients developed LVEF<50% and often had concurrent AF. Given this association and the morbidity associated with AF, aggressive rhythm management may be appropriate. Further studies are warranted to better characterize the long-term incidence of AF and LVSD with mavacamten therapy, and mechanistic investigations are needed to determine whether mavacamten directly influences AF burden.

## Data Availability

All data produced in the present study are available upon reasonable request to the authors.

## Abbreviations

AF: atrial fibrillation
CI: confidence interval
HCM: hypertrophic cardiomyopathy
HF: heart failure
LVEF: left ventricular ejection fraction
LVOT: left ventricular outflow tract
LVSD: left ventricular systolic dysfunction
oHCM: obstructive HCM
RCTs: randomized clinical trials
SCD: sudden cardiac death
SD: standard deviation

## References

1. Ho CY, Day SM, Ashley EA, et al. Genotype and Lifetime Burden of Disease in Hypertrophic Cardiomyopathy: Insights from the Sarcomeric Human Cardiomyopathy Registry (SHaRe). Circulation. 2018;138(14):1387–1398. doi:10.1161/CIRCULATIONAHA.117.033200

2. Siontis KC, Geske JB, Ong K, Nishimura RA, Ommen SR, Gersh BJ. Atrial Fibrillation in Hypertrophic Cardiomyopathy: Prevalence, Clinical Correlations, and Mortality in a Large HighCRisk Population. J Am Heart Assoc. 2014;3(3):e001002. doi:10.1161/JAHA.114.001002

3. Rowin EJ, Link MS, Maron MS, Maron BJ. Evolving Contemporary Management of Atrial Fibrillation in Hypertrophic Cardiomyopathy. Circulation. 2023;148(22):1797–1811. doi:10.1161/CIRCULATIONAHA.123.065037

4. Olivotto I, Oreziak A, Barriales-Villa R, et al. Mavacamten for treatment of symptomatic obstructive hypertrophic cardiomyopathy (EXPLORER-HCM): a randomised, double-blind, placebo-controlled, phase 3 trial. Lancet. 2020;396(10253):759–769. doi:10.1016/S0140-6736(20)31792-X

5. Davis BJ, Volk H, Nguyen O, et al. Safety and Efficacy of Mavacamten and Aficamten in Patients With Hypertrophic Cardiomyopathy. Journal of the American Heart Association. 2025;14(6):e038758. doi:10.1161/JAHA.124.038758

6. Boyle TA, Reza N, Hyman M, et al. Atrial Fibrillation in Patients Receiving Mavacamten for Obstructive Hypertrophic Cardiomyopathy: Real-World Incidence, Management, and Outcomes. JACC Clin Electrophysiol. 2025;11(2):411–413. doi:10.1016/j.jacep.2024.10.014

7. Castrichini M, Alsidawi S, Geske JB, et al. Incidence of newly recognized atrial fibrillation in patients with obstructive hypertrophic cardiomyopathy treated with Mavacamten. Heart Rhythm. 2024;21(10):2065–2067. doi:10.1016/j.hrthm.2024.04.055

8. Garcia-Pavia P, Oręziak A, Masri A, et al. Long-term effect of mavacamten in obstructive hypertrophic cardiomyopathy. European Heart Journal. 2024;45(47):5071–5083. doi:10.1093/eurheartj/ehae579

9. Desai MY, Wolski K, Owens A, et al. Mavacamten in Patients With Hypertrophic Cardiomyopathy Referred for Septal Reduction: Week 128 Results From VALOR-HCM. Circulation. 2025;151(19):1378–1390. doi:10.1161/CIRCULATIONAHA.124.072445

10. Desai MY, Owens AT, Abraham T, et al. Mavacamten in Symptomatic Nonobstructive Hypertrophic Cardiomyopathy. New England Journal of Medicine. 2025;393(10):961–972. doi:10.1056/NEJMoa2505927

11. Ferrantini C, Scellini B, Vitale G, et al. Mavacamten depresses human atrial contractility in the same EC50% range as human ventricle. Biophysical Journal. 2022;121(3):106a–107a. doi:10.1016/j.bpj.2021.11.2179

12. United States Food and Drug Administration Center for Drug Evaluation and Research. Clinical and Statistical Review(s): Mavacamten (Camzyos).; 2022. Accessed June 19, 2025. https://www.accessdata.fda.gov/drugsatfda_docs/nda/2022/214998Orig1s000Med_StatR.pdf

